# Genome-wide Polygenic Risk Scores Predict Risk of Glioma and Molecular Subtypes

**DOI:** 10.1101/2024.01.10.24301112

**Authors:** Taishi Nakase, Geno Guerra, Quinn T. Ostrom, Tian Ge, Beatrice Melin, Margaret Wrensch, John K. Wiencke, Robert B. Jenkins, Jeanette E. Eckel-Passow, Glioma International Case-Control Study (GICC), Melissa L. Bondy, Stephen S. Francis, Linda Kachuri

## Abstract

**Background:** Polygenic risk scores (PRS) aggregate the contribution of many risk variants to provide a personalized genetic susceptibility profile. Since sample sizes of glioma genome-wide association studies (GWAS) remain modest, there is a need to find efficient ways of capturing genetic risk factors using available germline data.

**Methods:** We developed a novel PRS (PRS-CS) that uses continuous shrinkage priors to model the joint effects of over 1 million polymorphisms on disease risk and compared it to an approach (PRS-CT) that selects a limited set of independent variants that reach genome-wide significance (P<5×10^-8^). PRS models were trained using GWAS results stratified by histological (10,346 cases, 14,687 controls) and molecular subtype (2,632 cases, 2,445 controls), and validated in two independent cohorts.

**Results:** PRS-CS was consistently more predictive than PRS-CT across glioma subtypes with an average increase in explained variance (R^2^) of 21%. Improvements were particularly pronounced for glioblastoma tumors, with PRS-CS yielding larger effect sizes (odds ratio (OR)=1.93, P=2.0×10^-54^ vs. OR=1.83, P=9.4×10^-50^) and higher explained variance (R^2^=2.82% vs. R^2^=2.56%). Individuals in the 95^th^ percentile of the PRS-CS distribution had a 3-fold higher lifetime absolute risk of *IDH* mutant (0.63%) and *IDH* wildtype (0.76%) glioma relative to individuals with average PRS. PRS-CS also showed high classification accuracy for *IDH* mutation status among cases (AUC=0.895).

**Conclusions:** Our novel genome-wide PRS may improve the identification of high-risk individuals and help distinguish between prognostic glioma subtypes, increasing the potential clinical utility of germline genetics in glioma patient management.

**IMPORTANCE OF THE STUDY:** Inherited genetic variation is one of only a few risk factors known to contribute to gliomagenesis. We leverage the largest available collection of genome-wide association studies for glioma to show that a genome-wide PRS approach that models the joint effect of correlated variants across the genome yields improved prediction of glioma risk. Our novel PRS also improves the classification of cases according to *IDH* mutation status. Additionally, we provide refined estimates of individual genetic susceptibility and show that risk scores in the highest percentiles of the PRS distribution confer significant increases in relative and lifetime absolute risk. Taken together, our findings provide further evidence of the potential for germline genotyping to be used as a clinical biomarker in the assessment of personalized glioma risk and the non-invasive management of patients with newly diagnosed brain tumors.

## INTRODUCTION

Adult diffuse glioma accounts for approximately 80% of all primary malignant central nervous system tumors with over 20,000 new cases each year in the United States^1^. These tumors encompass a highly heterogeneous group of subtypes with distinct genetic risk loci, molecular signatures, histopathologic lineages, etiological mechanisms and survival trajectories^2^. In particular, molecular tumor features including mutations in *IDH1* or *IDH2* (encoding the cytoplasmic and mitochondrial isocitrate dehydrogenase respectively), co-deletion of chromosome arms 1p and 19q, and mutations in *TERT* are used to define clinically relevant glioma subtypes^3^. Tumors without *IDH* mutation are primary glioblastomas (GBM) and carry the poorest prognosis with less than 5% of patients surviving more than 5 years^4^. Low grade astrocytomas are defined by *IDH* mutation without 1p19q codeletion and tend to have a better prognosis. Oligodendroglioma tumors, characterized by *IDH* mutation and 1p19q codeletion, carry the best prognosis with a median overall survival of 17.5 years^2^.

While the etiology of glioma remains poorly understood, multiple studies have shown that inherited genetic variation contributes to glioma susceptibility^5–7^. Genome-wide association studies (GWAS) of glioma patients with tumors stratified by histological subtype have identified approximately 25 common single nucleotide polymorphisms (SNPs) associated with glioma risk^7^. More recent glioma GWAS by molecular subtype have identified additional variants associated with *IDH*-mutant glioma in *D2HGDH* on chromosome 2 and *FAM20C* on chromosome 7^8^. Recognition that heritable genetic variants influence glioma risk has prompted the development of polygenic risk scores (PRS) to help identify high-risk individuals^9,10^. As a rare cancer glioma is not amenable to population-level screening, but a PRS that provides more accurate and personalized risk estimates, beyond family history and demographic factors, may be useful in certain scenarios such as refining a differential diagnosis. Additionally, predicting the risk of specific subtypes may help guide treatment and follow-up of patients with indeterminate brain lesions prior to invasive interventions^9^.

Previous PRS for glioma have used an approach known as clumping and thresholding (CT) which involves selecting a single GWAS-significant variant per linkage disequilibrium (LD) block^9,10^. These efforts have shown promising results with PRS greatly improving glioma risk prediction beyond demographic risk factors such as age and sex. However, since glioma is relatively rare, GWAS sample sizes, especially for contemporary glioma subtypes, remain modest with few variants reaching the conventional genome-wide significance threshold (P<5×10^-8^)^7,8^. As a result, the standard CT approach leaves a substantial component of the genetic liability for glioma unexplained^11^ and may discard information from variants below the significance threshold that could improve prediction accuracy. In contrast, genome-wide PRS approaches directly model the genetic architecture of the trait across the genome, using shrinkage methods to assign weights to each SNP according to their estimated contribution to the trait^12^. They have been shown to improve prediction performance compared to the CT method for several complex traits including schizophrenia^13^, major depressive disorder^13^, coronary artery disease^14^, breast cancer^14^ and colorectal cancer^15^. With small samples sizes yielding few GWAS-significant variants for glioma, PRS approaches that leverage information from variants that do not reach genome-wide significance thresholds may improve risk prediction across glioma subtypes.

In this investigation, we developed a novel PRS for glioma using a genome-wide Bayesian approach and assessed its ability to predict individual glioma risk and distinguish glioma subtypes compared to a standard CT PRS. Using our genome-wide PRS, we estimated lifetime absolute risk and cumulative incidence trajectories across histological and molecular subtypes.

## METHODS

### Glioma study population

An overview of the study design and analysis is provided in **Figure 1**. PRS models by histological subtype were trained using summary-level statistics from a meta-analysis of six studies provided by the Glioma International Case-Control (GICC) Consortium, which included all glioma (10,346 cases, 14,687 controls), GBM (5,395 cases, 14,687 controls) and non-GBM (4,466 cases, 14,687 controls) tumors (**Supplementary Table 1**), as described by Melin et al^7^. This GWAS was performed prior to the widespread adoption of molecular classifications such that the case definitions for GBM and non-GBM do not follow the current WHO 2021 guidelines^3^. Additionally, we leveraged data from two case-control studies to develop PRS for molecular subtypes of glioma: (1) the Mayo Clinic and University of California San Francisco (UCSF) Adult Glioma Study (1,973 cases) with controls from the GICC study (1,859 controls) and (2) UCSF Adult Glioma Study (659 cases, 586 controls)^8,9,16^. The study-specific summary statistics were meta-analyzed by *IDH* mutation status for a total sample size of 2,632 cases (1115 *IDH* wildtype, 699 *IDH* mutant, 818 *IDH* unknown) and 2,445 controls. Participants in the GWAS by molecular subtypes as well as those included in the individual-level genotype data used for testing were excluded from the GWAS meta-analysis where only histological subtypes were available. All analyses were restricted to individuals of predominantly European ancestry.

**Figure 1:**
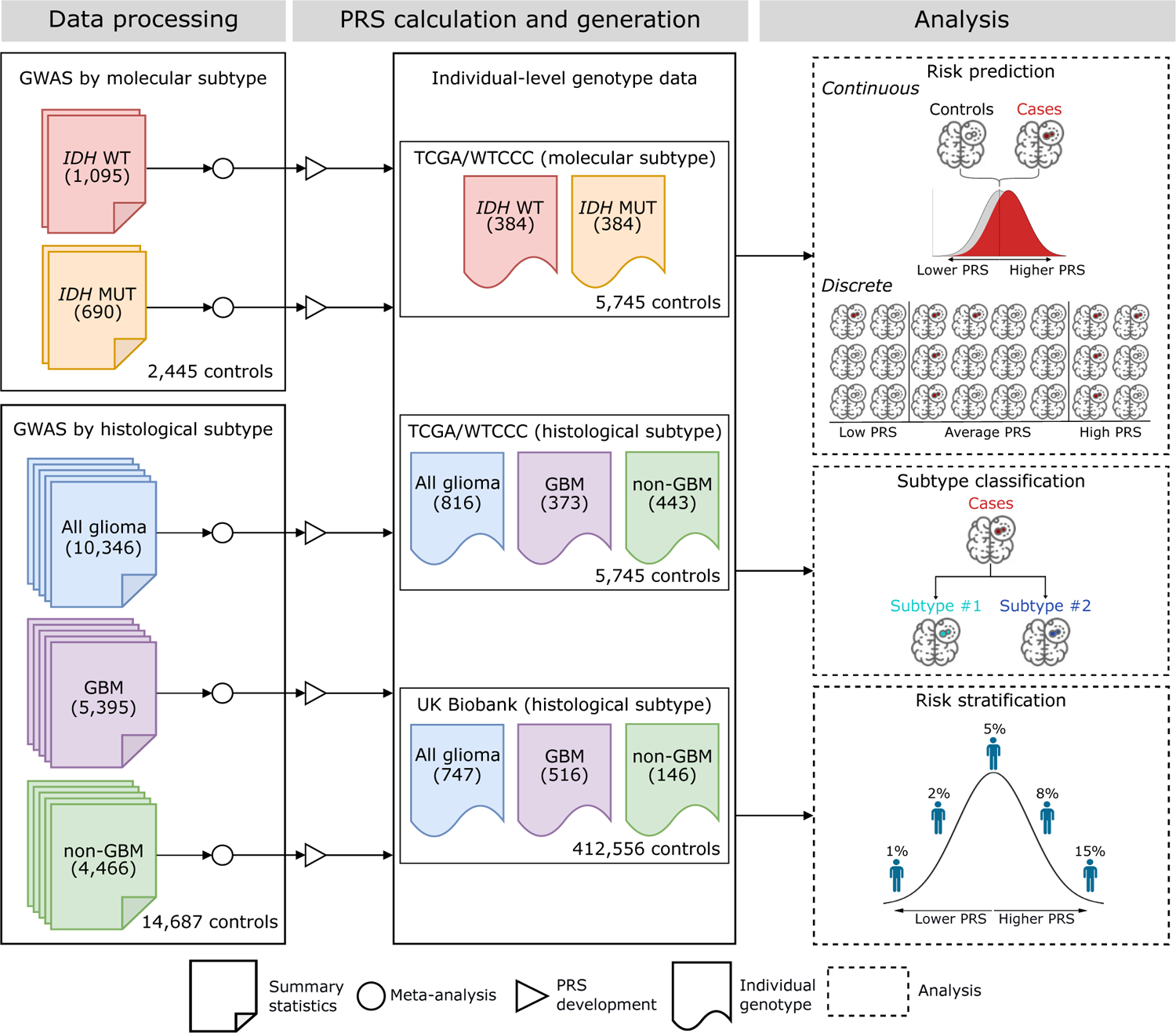
Overview of the study design.

The prediction accuracy of PRS models were tested in the UK Biobank (UKB)^17,18^, a population-based cohort with 747 glioma cases (516 GBM, 146 non-GBM) and 412,556 controls. PRS models for molecular subtypes were tested in The Cancer Genome Atlas (TCGA) with controls from the Wellcome Trust Case Control Consortium (WTCCC), as described in Guerra et al^19^. The TCGA/WTCCC dataset consisted of 814 glioma cases (372 GBM, 442 non-GBM; 384 *IDH* wildtype, 384 *IDH* mutant) and 5,745 controls.

### Polygenic risk score development and evaluation

We compared two different approaches to PRS construction: LD clumping and thresholding (PRS-CT)^20^ and a Bayesian genome-wide approach using continuous shrinkage priors (PRS-CS)^21^. For PRS-CT we preferentially selected variants with the lowest p-value from those with p<5×10^-8^ that were LD-independent (r2<0.05) within 10 Mb blocks (**Supplementary Table 2**). Each selected variant in PRS-CT was weighted by the marginal GWAS effect size. PRS-CS is an approach that uses penalized regression to shrink effect size estimates of variants across the genome according to the genetic architecture of the trait^21^. We expanded the panel of genetic markers used in PRS-CS to ensure adequate coverage of known glioma risk loci for a total of 1,120,700 variants present in HapMap3^22^.

Each PRS was converted to a standardized z-score based on the distribution in controls. Odds ratios (OR) per standard deviation (SD) increase in the PRS were estimated using logistic regression with adjustment for age (except for TCGA/WTCCC as controls were from the 1958 birth cohort), sex, and the top ten genetic ancestry principal components (PCs). The prediction accuracy of each PRS was quantified using two metrics: pseudo-R^2^ on the liability scale^23^ and the area under the receiver operator characteristic curve (AUC). The relative increase or decrease in R^2^ of PRS-CS compared to PRS-CT is defined as (R^2^_CS_-R^2^_CT_)/R^2^_CT_.

In addition to case/control analyses we also evaluated the potential of PRS to classify glioma cases according to *IDH* mutation status in the TCGA testing dataset. Models that included PRS-CT or PRS-CS were compared to a baseline model with age and sex. Model discrimination was assessed using AUC with 95% confidence intervals estimated from 2,000 bootstrap samples.

### Assessment of risk stratification

We evaluated the risk stratification potential of PRS by estimating age-specific incidence trajectories for glioma cases in UKB, following the approach in Kachuri et al^17^. Cumulative incidence estimates were obtained from cause-specific Cox regression models for incident glioma cases with all-cause mortality treated as a competing event^24,25^. Cox models were adjusted for age, sex and the first ten genetic ancestry PCs. Cumulative incidence trajectories were compared across PRS strata with individuals above the 80^th^ percentile of the PRS distribution defined as having high genetic risk, those below the 20^th^ percentile as low risk and those in the middle 20^th^-80^th^ percentile as average risk.

Additionally, we compared the lifetime absolute risk of developing glioma for individuals across PRS levels using the method described by Pain et al^26^. Briefly, the approach defines the population distribution of the risk score within normal distribution theory using the estimated OR per SD unit increase in PRS and then calculates the proportion of cases within risk score quantiles. Lifetime risk of each glioma subtype in the general population was obtained from the Surveillance, Epidemiology and End Results (SEER) Program (https://seer.cancer.gov/). Confidence intervals for absolute risk estimates were obtained by parametric bootstrap sampling of the OR estimates of each PRS and each glioma subtype.

## RESULTS

### Comparison of PRS approaches for predicting glioma risk

PRS-CS generally showed improved performance over PRS-CT across all glioma subtypes except non-GBM tumors with consistent results in the TCGA/WTCCC and UKB testing datasets (**Figure 2**). For glioma overall, PRS-CS showed a larger magnitude of association (OR per SD = 1.53, 95% CI: 1.40-1.69, P=1.1×10^-18^) than PRS-CT (OR=1.44, 95% CI: 1.31-1.58, P=4.5×10^-14^) in TCGA/WTCCC (**Supplementary Table 3**). The same pattern was observed in UKB, although the difference between the two approaches was attenuated (PRS-CS: OR=1.70, P=5.7×10^-54^; PRS-CT: OR=1.64, P=4.1×10^-49^). Additionally, PRS-CS achieved an increase in R^2^ of 23.6% and 11.0% relative to PRS-CT for TCGA/WTCCC and UKB, respectively (**Supplementary Table 3**).

**Figure 2:**
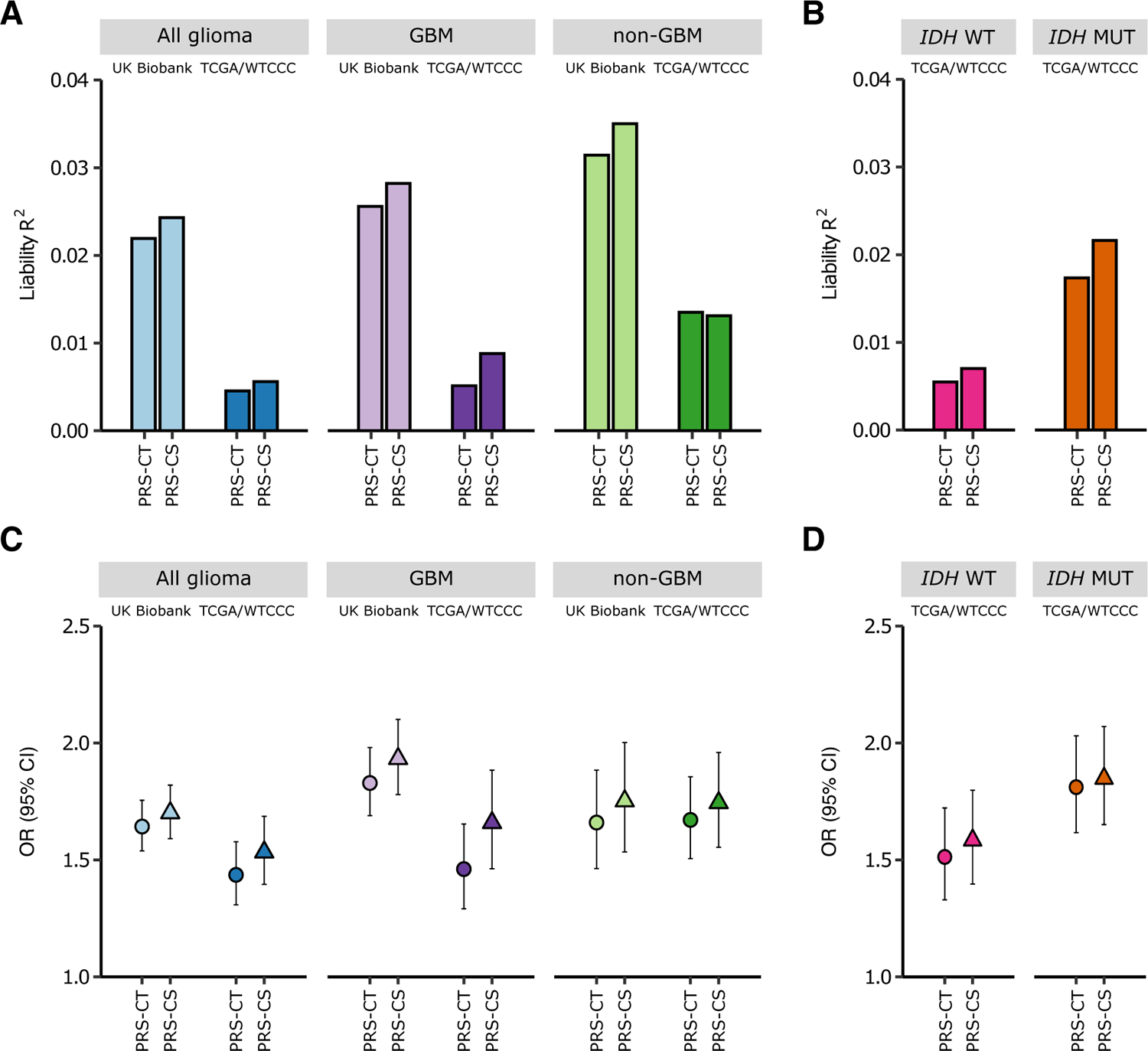
Prediction accuracy of PRS-CT and PRS-CS in the UK Biobank and TCGA/WTCCC datasets across glioma subtypes. (**A-B**) Prediction accuracy, measured as variance explained (R^2^) on the liability scale, for PRS trained using discovery GWAS summary statistics specific to target phenotype (histological: all glioma, GBM, non-GBM; molecular: *IDH* wildtype, *IDH* mutant). Each bar represents one testing cohort with the shade of the bar representing the testing dataset (light=UK Biobank; dark=TCGA/WTCCC). (**C-D**) The odds ratio (OR) per standard deviation (SD) unit increase in PRS for each glioma subtype in the UK Biobank and TCGA/WTCCC testing datasets. Each dot corresponds to one testing cohort with the shape of the dot representing the PRS construction method (circle=PRS-CT; triangle=PRS-CS) and the shade of the dot representing the testing dataset (light=UK Biobank; dark=TCGA/WTCCC). The error bars represent the 95% confidence intervals (CIs). The R^2^, ORs and 95% CIs for each subtype, PRS construction method and testing cohort are provided in Supplementary Tables 3-5.

In TCGA/WTCCC, PRS-CS (OR=1.66, 95% CI: 1.46-1.88, P=5.6×10^-15^) was more predictive of GBM than PRS-CT (OR=1.46, 95% CI: 1.29-1.65, P=2.2×10^-9^). The ORs for GBM risk were also higher for PRS-CS (OR=1.93, 95% CI: 1.78-2.10, P=2.0×10^-54^) than PRS-CT (OR=1.83, 95% CI: 1.69-1.98, P=9.4×10^-50^) in UKB. Similar improvements were obtained in terms of R^2^ (TCGA/WTCCC: 71.2% increase; UKB: 10.2% increase).

For non-GBM tumors, both methods showed comparable results (**Supplementary Table 3**). In UKB, PRS-CS demonstrated a larger effect size (OR=1.75, P=1.7×10^-16^) and modestly improved discrimination (AUC=0.676) over PRS-CT (OR = 1.66, P=4.6×10^-15^, AUC=0.667). In contrast, we observed slightly higher explained variance for PRS-CT (R^2^=0.0135) than PRS-CS (R^2^=0.0131) in TCGA/WTCCC. Given the heterogeneity among the histopathological lineages of non-GBM tumors, we examined each subtype encompassed by the non-GBM category separately (**Supplementary Table 4**). Both genetic scores were less predictive of astrocytoma risk (PRS-CS: OR=1.43, P=9.2×10^-5^; PRC-CT: OR=1.41, P=6.8×10^-5^), but showed stronger associations with oligodendroglioma (PRS-CS: OR=2.18, P=5.2×10^-11^; PRS-CT: OR=1.92, P=5.5×10^-9^) in UKB. Similar results were observed in TCGA/WTCCC.

Next we sought to examine how well a PRS trained on GWAS summary statistics for GBM and non-GBM tumors predicts risk of *IDH* wildtype and *IDH* mutant tumors respectively in TCGA/WTCCC (**Supplementary Table 5**). For *IDH* wildtype tumors, PRS-CS trained on GBM GWAS results (OR=1.72, P=2.3×10^-17^, R^2^=0.0107) showed marked improvement over PRS-CT (OR=1.51, P=4.2×10^-11^, R^2^=0.0063). For *IDH* mutant glioma, PRS-CS did not show improved performance over PRS-CT when trained using non-GBM GWAS results. The advantage of the genome-wide PRS-CS approach became more pronounced when models were trained on the smaller molecular-based GWAS that yielded fewer GWAS significant variants (**Figure 2**, **Supplementary Table 5**). For *IDH* wildtype gliomas PRS-CS showed a larger effect size (OR=1.58, P=1.0×10^-12^) and a 28% increase in explained variance compared to PRS-CT (OR=1.51, P=4.0×10^-10^). The difference between the two approaches was slightly smaller for *IDH* mutant gliomas (PRS-CS: OR=1.85, P=2.5×10^-26^; PRS-CT: OR=1.81, P=2.5×10^-24^).

Lastly, we estimated associations across percentiles of PRS to examine the shape of the risk gradient. Changes in risk were not monotonic with individuals above the 80^th^ and 95^th^ percentiles showing substantially elevated risk relative to those between the 40^th^ and 60^th^ percentiles (**Figure 3**, **Supplementary Table 6**). PRS-CS generally showed larger effect sizes than PRS-CT, especially for molecular subtypes of glioma (**Supplementary** Figure 1). Individuals above the 95^th^ percentile of PRS-CS had a significantly higher risk of *IDH* wildtype glioma (OR=2.85, 95% CI: 1.74-4.66) and *IDH* mutant glioma (OR=8.05, 95% Ci: 4.86-13.31) relative to those with average genetic predisposition.

**Figure 3:**
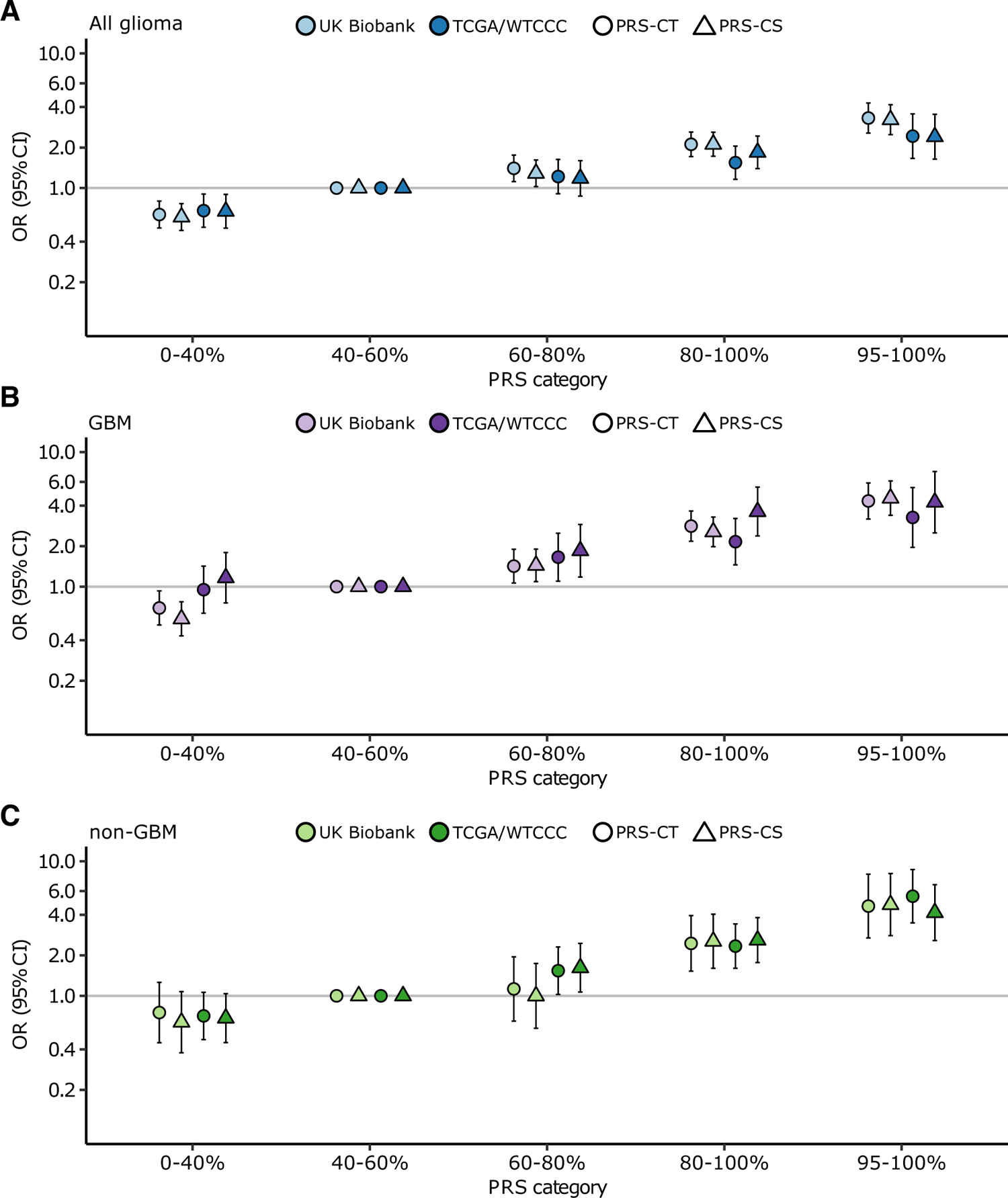
Relative glioma risk by PRS category stratified by histological subtype. (**A**) All glioma. (**B**) GBM subtype. (**C**) Non-GBM subtype. The x axis indicates the percentiles of the PRS distribution (0-40%, 40-60%, 60-80%, 80-100%, 95-100%). The y axis indicates odds ratios (ORs) with error bars representing 95% confidence intervals (CIs) for each PRS category relative to the middle category (40-60%) of risk scores. The results are stratified by testing dataset (light=UK Biobank; dark=TCGA/WTCCC) and PRS method (circle=PRS-CT; triangle=PRS-CS). Each PRS (colored shape) is trained using discovery GWAS summary statistics that correspond to the target phenotype.

### Using PRS to predict IDH status among glioma cases

In addition to evaluating the ability of PRS to discriminate between glioma cases and cancer-free controls, we examined the potential of PRS to distinguish glioma cases with and without *IDH* mutations in TCGA (384 *IDH* mutant; 384 *IDH* wildtype). First, we evaluated risk scores trained on GWAS results stratified by histological subtype (**Figure 4**, **Supplementary Table 7**). Without demographic covariates, the non-GBM PRS (PRS-CS: AUC=0.691, 95% CI: 0.653-0.726; PRS-CT: AUC=0.713, 95% CI: 0.679-0.750) showed improved subtype discrimination compared to the GBM PRS (PRS-CS: AUC=0.674, 95% CI: 0.636-0.711; PRS-CT: AUC=0.632, 95% CI: 0.592-0.669). Replacing GBM and non-GBM PRS with scores trained on IDH wildtype (PRS-CS: AUC=0.647; PRS-CT: AUC=0.630) and *IDH* mutant glioma (PRS-CS: AUC=0.714; PRS-CT: AUC=0.716) did not consistently increase classification accuracy.

**Figure 4:**
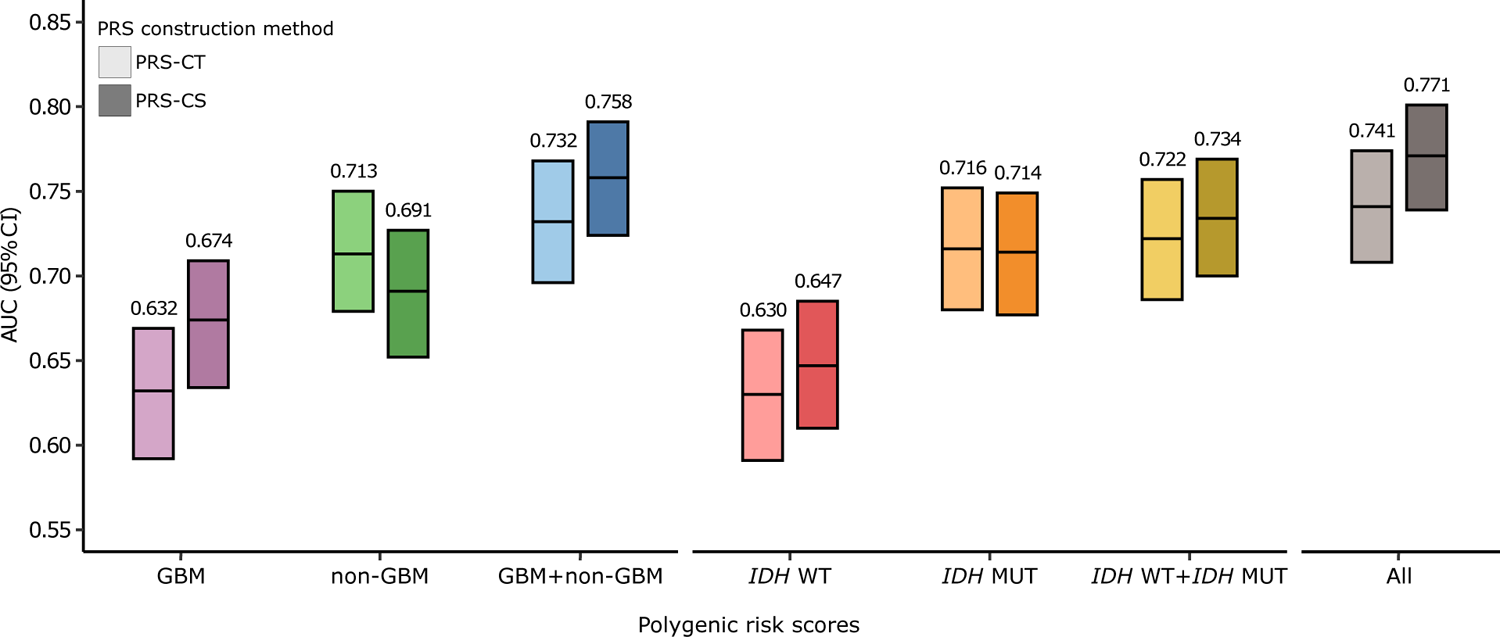
Comparison of PRS performance for logistic regression models that classify TCGA cases according to *IDH* mutation status. Classification accuracy of TCGA cases (384 *IDH* mutant; 384 *IDH* wildtype) for PRS-CT and PRS-CS using PRS trained on discovery GWAS summary statistics with histological (GBM or non-GBM) or molecular profiling (*IDH* mutant and *IDH* wildtype). Each bar represents the area under the receiver operating characteristic curve (AUC) and 95% confidence interval (CI) (2,000 bootstraps) for a single logistic regression model adjusted for the first 10 genetic ancestry principal components with the shade of the bar representing the PRS construction method (light=PRS-CT; dark=PRS-CS). The x axis indicates the PRS included in each model (single: GBM [purple], non-GBM [green], *IDH* wildtype [red], *IDH* mutant [orange]; multiple: GBM + non-GBM [blue], *IDH* mutant + *IDH* wildtype [yellow], all=GBM + non-GBM + *IDH* mutant + *IDH* wildtype [grey]). Results with additional covariates are provided in Supplementary Table 7.

The low correlation between the GBM PRS and the non-GBM PRS (Pearson *r*=0.27) suggested that these scores might contribute orthogonal information. Among PRS based on histological subtypes alone, the combination non-GBM and GBM PRS-CS achieved the best performance with an AUC of 0.758 (95% CI: 0.724-0.792). Similarly, combining PRS-CS for *IDH* wildtype and *IDH* mutant gliomas produced improved performance with an AUC of 0.734 (95% CI: 0.699-0.770). Since there was no overlap in participants included in the training GWAS for histology-based PRS (GBM and non-GBM) and molecular-based PRS (*IDH* wildtype and *IDH* mutant), we included both sets of scores in a single model. This approach achieved the best overall classification accuracy with an AUC of 0.771 (95% CI: 0.737-0.803) for PRS-CS and 0.741 (95% CI: 0.708-0.774) for PRS-CT (**Supplementary Table 7**). Incorporating age and sex increased the PRS-CS AUC to 0.895 (95% CI: 0.872-0.918).

### Incorporating PRS to improve risk stratification

Age-specific 5-year cumulative incidence trajectories diverged significantly between individuals with a high-risk PRS-CS profile (>80th percentile) compared to those with low (<20th percentile) and intermediate (>20th to <80th percentile) PRS profiles (**Figure 5**, **Supplementary** Figure 2). At age 60, risk increased from 0.11% to 0.23% for the high risk PRS profile in males and from 0.07% to 0.14% in females (**Supplementary Table 8**). For GBM tumors, PRS-CS also produced distinct cumulative incidence trajectories, with 5-year risk increasing from 0.07% (average PRS) to 0.16% (high PRS) in 60-year-old males and from 0.04% to 0.08% in 60-year-old females. The degree of risk stratification provided by PRS-CS was lower for non-GBM tumors, with overlapping age-specific incidence trajectories.

**Figure 5:**
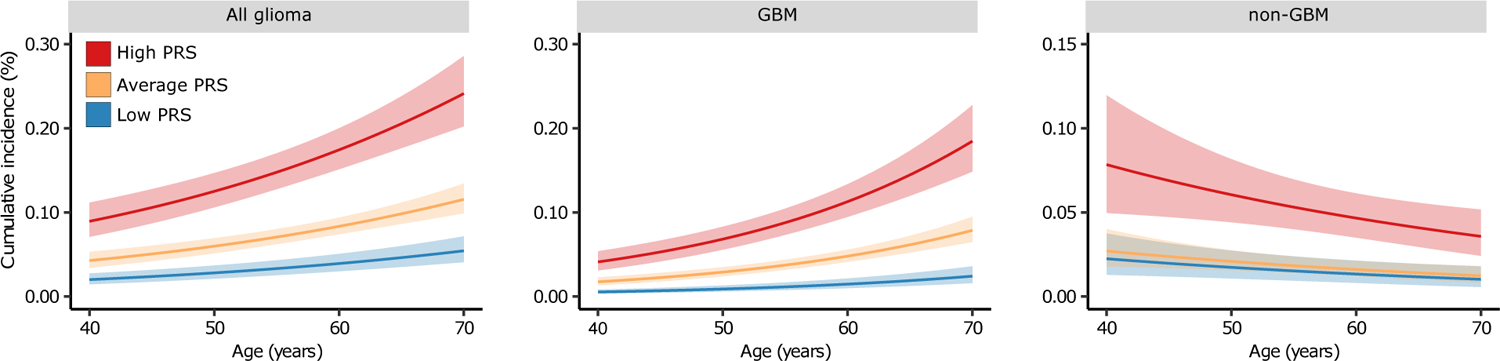
Estimated 5-year cumulative incidence as a function of age stratified by percentiles of the PRS-CS distribution for a typical individual of European ancestry in UK Biobank. Low PRS corresponds to below the 20^th^ percentile, average PRS is defined as between the 20^th^ and 80^th^ percentiles and high PRS includes individuals above the 80^th^ percentile of the normalized PRS-CS distribution. Each PRS-CS is trained on discovery GWAS summary statistics corresponding to the target phenotype (all glioma, GBM and non-GBM). The shaded areas represent 95% confidence intervals.

We found large differences in the lifetime absolute risk of developing glioma across the genetic susceptibility distribution estimated using PRS-CS (**Figure 6**, **Supplementary Table 9**). In UKB, the lifetime risk of glioma increased from 0.46% (95% CI: 0.45-0.48%) for average-risk individuals in the 40-60th percentiles to 1.39% (95% CI: 1.25-1.55%) for the highest-risk individuals above the 95^th^ percentile. The increase in risk was similar for GBM and non-GBM tumors. We also observed significant differences in the lifetime risk of *IDH* wildtype (0.29% vs. 0.76%) and *IDH* mutant gliomas (0.17% vs. 0.63%) between the average-risk and the highest-risk individuals.

**Figure 6:**
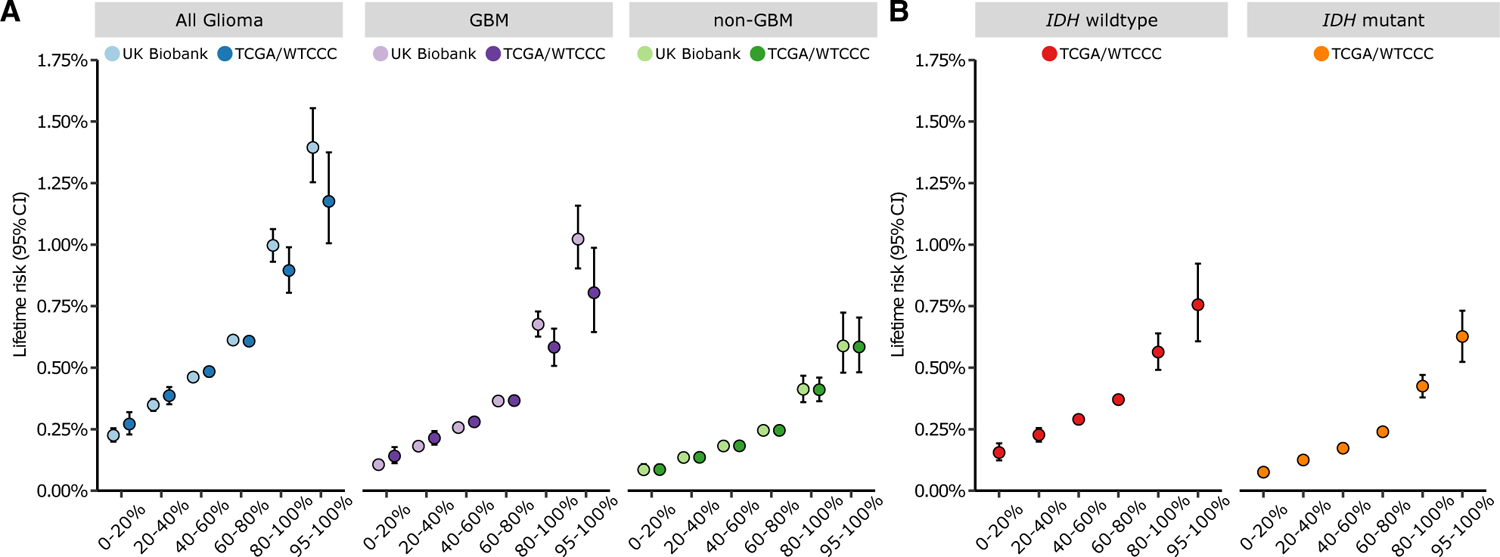
Lifetime absolute risks for each glioma subtype by PRS category. (**A**) Histological subtypes (all glioma, GBM and non-GBM). (**B**) Molecular subtypes (*IDH* wildtype and *IDH* mutant). The x axis indicates the PRS percentile, and the y axis is the lifetime absolute risk with error bars for the 95% confidence intervals (Cis). Each PRS (colored dots) is trained on discovery GWAS summary statistics that correspond to the target phenotype (labeled grey bar) using PRS-CS with the shading of the dots representing the testing cohort (light=UK Biobank; dark=TCGA/WTCCC).

## DISCUSSION

In this study we developed a novel PRS for glioma and its molecular subtypes using a genome-wide Bayesian approach with a custom reference panel designed to robustly tag known glioma risk loci. Previous PRS efforts for glioma have implemented variations of the standard clumping and thresholding approach, constructing risk scores based on a limited number of variants that reach genome-wide significance thresholds^9,10^. Our findings suggest that PRS approaches that model the joint effects of correlated variants across the genome may capture the genetic architecture of glioma more comprehensively than approaches that select independent risk variants from GWAS-identified risk loci. These results are consistent with observations for several complex traits, including other cancers^13–15^. We found that PRS-CS had improved prediction accuracy in terms of R^2^ and AUC, and higher effect sizes compared to PRS-CT for glioma overall, GBM, *IDH* mutant tumors and *IDH* wildtype tumors. However, PRS-CS did not show an advantage for non-GBM gliomas, possibly due to greater within-subtype heterogeneity which may limit the ability of PRS-CS to accuracy identify truly non-zero signals^21^.

We also found that the genome-wide PRS approach was highly effective at classifying gliomas according to *IDH* mutation status (AUC=0.895). Interestingly, PRS models trained on *IDH* wildtype and *IDH* mutant GWAS results did not outperform PRS developed based on GWAS stratified by histological labels alone. However, the GBM and non-GBM PRS were trained on a substantially larger GWAS than the molecular-based PRS. Since most GBM and non-GBM cases were likely *IDH* wildtype and *IDH* mutant, respectively, the larger sample size may compensate for some misclassification when using case definitions based on histology alone to predict *IDH* mutation status. This finding should be interpreted with caution since a high degree of phenotype misclassification can lead to dilution and decrease PRS accuracy^27^. Increasing GWAS sample sizes for gliomas defined using contemporary molecular classifications thus remains important for improving the accuracy of genetic prediction models. Importantly, we also showed that combining risk scores developed using histological and molecular GWAS data is a viable strategy that significantly improves discrimination of *IDH* mutation status.

In addition to disease and subtype classification, we also evaluated the potential of PRS to refine risk stratification. We found that the lifetime absolute risk for individuals with the highest inherited genetic risk was significantly greater than that for individuals with average genetic risk profiles. The low lifetime absolute risk of glioma even in individuals with the highest genetic risk, however, presents a barrier to the use of PRS for population-level disease screening since disease risk for most screened individuals would be too low to warrant an intervention. In the first study of genetic scores for glioma subtypes, Eckel-Passow^9^ suggests that PRS could instead be used in conjunction with diagnostic imaging to characterize suspicious brain lesions. For example, a glioma risk score could help distinguish among high-grade glioma, solitary metastases^28,29^, meningiomas^29^, primary central nervous system lymphomas^28^ and tumefactive demyelinating lesions^30^ which often display similar imaging features but require substantially different management. Since the differentiation of brain lesions remains a challenge, risk scores that identify individuals with a genetic predisposition to glioma could provide useful clinical context for the interpretation of indeterminate imaging findings and help refine the differential diagnosis.

Molecular markers, together with tumor morphology, have been shown to accurately predict prognosis^31–33^ and treatment response^34^. Since establishing a molecular diagnosis early is important for prediction of tumor behavior and clinical management^35,36^, tumor molecular features such as *IDH* mutation have been explored as potential targets for cancer therapeutics. Vorasidenib, an inhibitor of mutant *IDH1* and *IDH2* enzymes, was recently shown in a phase 3 trial to improve progression-free survival and delay the time to next intervention in patients with grade 2 *IDH*-mutant glioma^35^, becoming the first therapy targeted to a specific molecular subtype. Currently, the classification of brain tumors is based on immunohistochemical and genomic sequence analysis of tumor specimens, which often delays the evaluation of patient prognosis and treatment options until after surgery. Given the invasiveness of tumor resection and the risks of delaying treatment until after surgery, research has been directed at the use of non-invasive procedures such as imaging^37–39^ and genotyping^9^ to classify glial tumors prior to surgery. Our study provides further support for the potential use of germline genetic PRS, which can be easily calculated, to help predict *IDH* mutation status in gliomas. Importantly, our results add to the collection of non-invasive tools that could be used to help select patients suitable for neoadjuvant therapy with *IDH* inhibitors^35^, assist in preoperative planning of extent of resection^40^ and accurately delineate *IDH* wildtype non-enhancing tumors which often display characteristics of low-grade tumors on imaging but require aggressive treatment.

In evaluating this study, several limitations should be acknowledged. Our TCGA/WTCCC validation dataset used external controls, which may introduce bias and inflate type I error due to batch effects and inadequate data harmonization. We attempted to limit the influence of technical artifacts by selecting WTCCC controls that were genotyped using the same array as TCGA cases as well as conducting systematic genotype harmonization and rigorous pre- and post-imputation quality control. We also used the UK Biobank, a population-based cohort, as an additional validation dataset. Although the UK Biobank lacked information on molecular subtypes, we observed consistent and robust results for PRS analyses of glioma overall and histological subtypes. Additionally, prediction performance for non-GBM tumors may have been limited by differences in the relative proportions of different histopathologic lineages comprising non-GBM between the GWAS training and PRS testing datasets. As larger studies comprised of tumors specimens with more refined molecular classification become available, the accuracy and utility of PRS models for glioma are expected to improve. Finally, our study was limited to participants of European ancestry and the developed PRS models are expected to have lower accuracy in populations with diverse and predominantly non-European ancestral backgrounds^41,42^. Future research efforts should prioritize recruitment of more diverse patients populations to help elucidate the genetic underpinnings of glioma and increase the clinical utility of genetic prediction models.

This work has several important strengths. Our study develops a novel PRS model for glioma and its subtypes that may better capture the underlying genetic architecture of glioma than previous models that were limited to a small number of genetic markers. Each PRS model is trained using the largest available collection of glioma GWAS data across glioma subtypes and tested in two independent cohorts. Furthermore, we introduce an expanded linkage disequilibrium reference panel for PRS-CS that is designed to more comprehensively characterize known glioma risk loci across the genome. The development of custom reference panels may be particularly advantageous for PRS modelling of other complex traits with similarly sparse genetic architecture that is not well characterized by commonly used reference panels. Finally, we use our novel PRS to provide a comprehensive description of the relative influence of genetic risk factors on the lifetime risk of developing glioma. Further improvements in risk stratification could be achieved by incorporating additional non-genetic risk factors such as family history and varicella-zoster virus (VZV) antibody seropositivity^43,44^ into integrated glioma risk models.

In summary, we found that PRS for glioma has predictive power for both general risk modeling and subtype delineation. Across histological and molecular subtypes of glioma, the genome-wide approach to PRS modeling consistently showed improved risk prediction, risk stratification and subtype classification.

## FUNDING

L.K. is supported by funding from the National Institutes of Health (NIH): R00CA246076. T.N. is supported by the Stanford School of Medicine, MedScholars Research Fellowship. Work at University of California, San Francisco was supported by the NIH (grant numbers T32CA151022, R01CA52689, P50CA097257, R01CA126831, R01CA139020, R01AI128775, and R25CA112355), the National Brain Tumor Foundation, the Stanley D. Lewis and Virginia S. Lewis Endowed Chair in Brain Tumor Research, the Robert Magnin Newman Endowed Chair in Neuro-oncology, and by donations from families and friends of John Berardi, Helen Glaser, Elvera Olsen, Raymond E. Cooper, and William Martinusen. This publication was supported by the National Center for Research Resources and the National Center for Advancing Translational Sciences, National Institutes of Health, through UCSF-CTSI grant number UL1 RR024131. The collection of cancer incidence data used in this study was supported by the California Department of Public Health pursuant to California Health and Safety Code Section 103885; Centers for Disease Control and Prevention’s (CDC) National Program of Cancer Registries, under cooperative agreement 5NU58DP006344; the National Cancer Institute’s Surveillance, Epidemiology, and End Results Program under contract HHSN261201800032I awarded to the University of California San Francisco, contract HHSN261201800015I awarded to the University of Southern California, and contract HHSN261201800009I awarded to the Public Health Institute, Cancer Registry of Greater California. Work at the Mayo Clinic was supported by NIH grants CA230712, P50 CA108961, and CA139020; the National Brain Tumor Society; the loglio Collective; the Mayo Clinic; and the Ting Tsung and Wei Fong Chao Foundation.

## Data Availability

Genotype data of glioma cases from The Cancer Genome Atlas (TCGA) are available from the Database of Genotypes and Phenotypes (dbGaP) under accession phs000178. Genotype data of control samples from the 1958 British Birth Cohort and UK Blood Service Control Group are provided by the Wellcome Trust Case Control Consortium (WTCCC) and can be downloaded from the European Genotype Archive under ascension numbers EGAD00000000021 and EGAD00000000023. This research has been conducted using the UK Biobank Resource under application number 14015. UK Biobank data are publicly available by request from https://www.ukbiobank.ac.uk. Meta-analyzed GWAS summary statistics were obtained from Melin et al^7^. Genotype data from the Glioma International Case-Control Study (GICC) GWAS are available from dbGaP under accession phs001319.v1.p1. Genotype data from the GliomaScan GWAS can be accessed through dbGaP under accession phs000652.v1.p1. Genotype data from the UCSF Adult Glioma Study are available from dbGaP under accession phs001497.v2.p1.

## ACKNOWLEDGEMENTS

We would like to thank John S. Witte for providing helpful feedback and discussions.

## CONFLICT OF INTEREST STATEMENT

The authors declare no competing interests.

## ETHICS STATEMENT

Collection of patient samples and associated clinicopathological information was undertaken with written informed consent and ethical board approval was obtained from the UCSF Committee on Human Research (USA) and the Mayo Clinic Office for Human Research Protection (USA).

## AUTHORSHIP STATEMENT

T.N and L.K. conceived the project. T.N., T.G. and L.K. advised on the methodology. T.N. performed the main analyses. T.N., G.G., Q.T.O, B.M., M.W., J.K.W., R.B.K, J.E.E.P., GICC, M.L.B., S.S.F., L.K. were involved in primary data collection and curation. T.N. and L.K. drafted the manuscript and all authors contributed to, reviewed, and approved the final manuscript.

## Notes

### Competing Interest Statement

The authors have declared no competing interest.

### Author Declarations

Ethics committee of the UCSF Committee on Human Research (USA) and the Mayo Clinic Office for Human Research Protection (USA) gave ethical approval for this work.

