## Supplementary Figures S1-S2,Supplementary Tables S1-S9 for "Genome-wide Polygenic Risk Scores Predict Risk of Glioma and Molecular Subtypes"

**Supplementary Figure S1: Relative glioma risk by PRS category stratified by histological subtype.** (**A**) *IDH* wildtype. (**B**) *IDH* mutant. The x axis indicates the percentiles of the PRS distribution (0-40%, 40-60%, 60-80%, 80-100%, 95-100%). The y axis indicates odds ratios (ORs) with error bars representing 95% confidence intervals (CIs) for each PRS category relative to the middle category (40-60%) of risk scores. The results are stratified by PRS method (circle=PRS-CT; triangle=PRS-CS) using the TCGA/WTCCC testing dataset. Each PRS (colored shape) is trained using discovery GWAS summary statistics that correspond to the target phenotype.

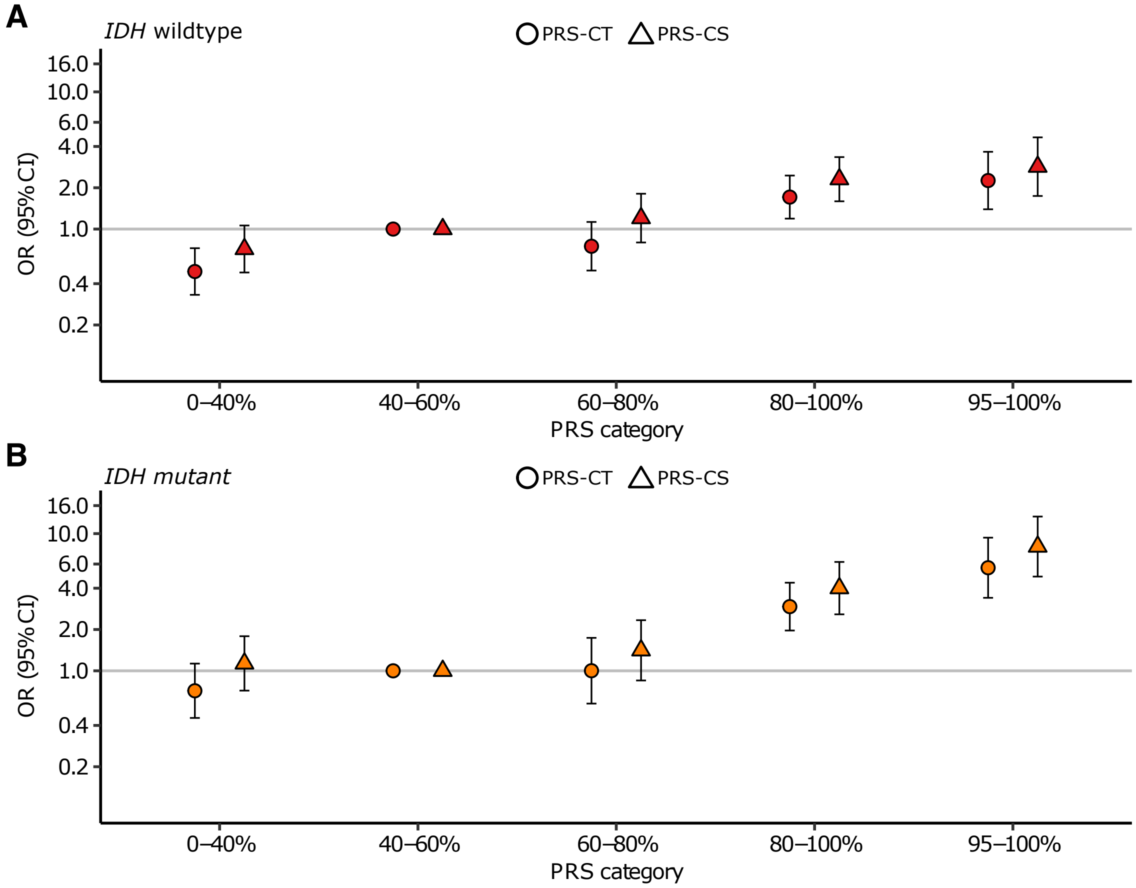

**Supplementary Figure 2: Estimated 5-year cumulative incidence as a function of age and sex stratified by percentiles of the polygenic risk score (PRS-CS) distribution for a typical individual of European ancestry in UKB.** Low PRS corresponds to below the 20th percentile, average PRS is defined as between the 20th and 80th percentile and high PRS includes individuals above the 80th percentile of the normalized PRS distributions. Each PRS-CS is trained on discovery GWAS summary statistics corresponding to the target phenotype (all glioma, GBM and non-GBM). The shaded areas represent 95% confidence intervals.

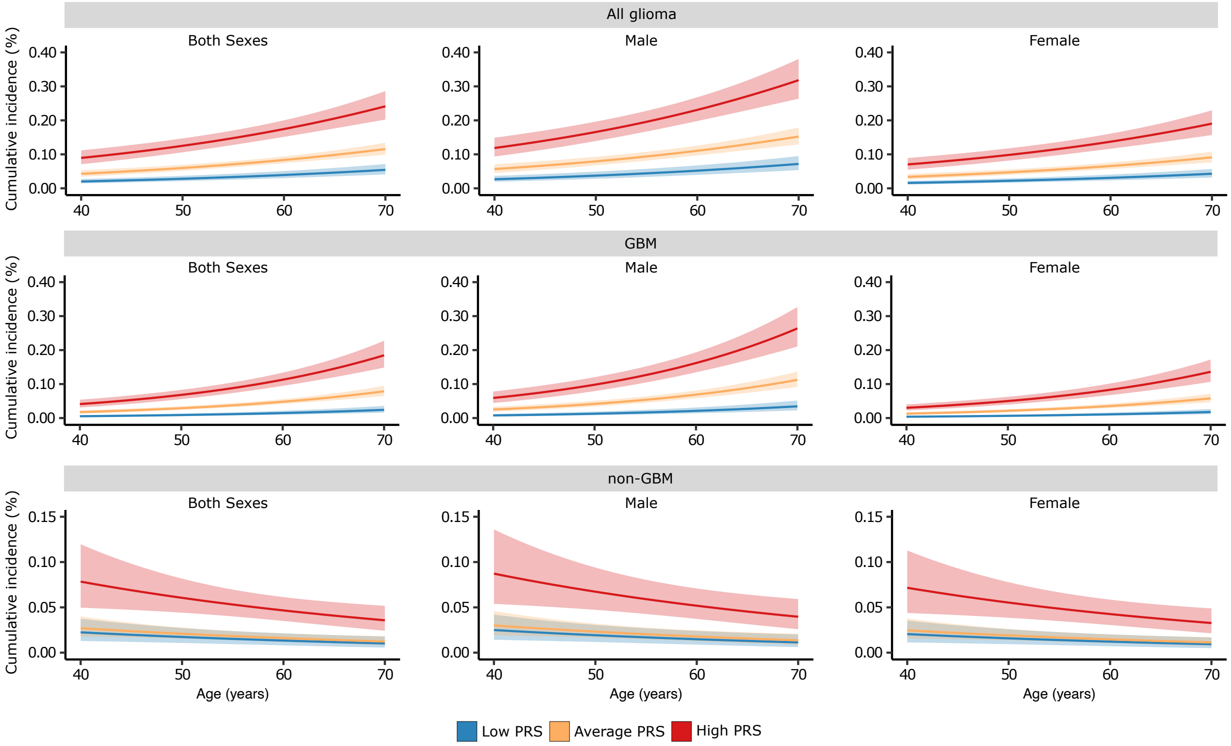

**Supplementary Table 1: Details of studies used to train PRS models by histological subtype.** Detailed information about individual studies including quality control are provided in Melin et al[7].

| Study | Cases (GBM/non-GBM) | Controls |
|---|---|---|
| French-GWAS | 1,423 (430/993) | 1,190 |
| German-GWAS | 846 (431/415) | 1,310 |
| MDA-GWAS | 1,175 (652/523) | 2,236 |
| SFAGS | 677 (511/166) | 3,940 |
| GliomaScan | 1,653 (903/472) | 2,725 |
| GICC | 4,572 (2,468/1,897) | 3,286 |
| Total | 10,346 (5,395/4,466) | 14,687 |

**Supplementary Table 2: Single nucleotide polymorphisms included in each of the polygenic risk scores using the PRS-CT approach.** Weights are only reported for SNPs that are used in the PRS for each glioma subtype. Weights are reported as log(odds ratio).

| Chr | Position (GRCh37) | rsID | Other Allele | Effect Allele | GWAS Effect Size | | | | |
|---|---|---|---|---|---|---|---|---|---|
| | | | | | All glioma | GBM | Non-GBM | IDH WT | IDH MUT |
| 1 | rs12752552 | 65229299 | C | T | 0.1747 | 0.2214 | - | - | - |
| 1 | rs3006933 | 243659727 | G | A | - | - | 0.1474 | - | - |
| 2 | rs7572263 | 209051586 | G | A | - | - | 0.1964 | - | - |
| 5 | rs4975605 | 1275528 | A | C | - | 0.1580 | - | - | - |
| 5 | rs7726159 | 1282319 | C | A | 0.3409 | - | - | 0.5339 | - |
| 5 | rs4449583 | 1284135 | C | T | - | - | 0.2076 | - | - |
| 5 | rs7705526 | 1285974 | C | A | - | 0.4379 | - | - | - |
| 5 | rs34363858 | 1286477 | G | T | - | 0.5133 | - | - | - |
| 5 | rs12332579 | 1311198 | T | C | 0.2082 | 0.2652 | - | - | - |
| 7 | rs75061358 | 54916280 | T | G | 0.3560 | 0.5116 | - | - | - |
| 7 | rs1015793 | 55114316 | G | A | - | 0.3151 | - | - | - |
| 7 | rs6960438 | 55141908 | T | C | 0.1847 | 0.2441 | - | - | - |
| 7 | rs17172438 | 55151537 | C | T | 0.2026 | - | - | - | - |
| 7 | rs7808726 | 55190206 | C | T | - | 0.6132 | - | - | - |
| 8 | rs12681076 | 130456426 | T | C | - | - | 0.1655 | - | - |
| 8 | rs55705857 | 130645692 | A | G | 0.5923 | - | 1.1531 | - | 1.5123 |
| 8 | rs4377928 | 130649867 | G | T | - | - | 0.1937 | - | - |
| 8 | rs6997969 | 130661870 | T | C | 0.1280 | - | - | - | - |
| 8 | rs62523827 | 130697170 | A | G | - | - | 0.2819 | - | - |
| 8 | rs142586905 | 130903744 | T | C | - | - | 0.5963 | - | - |
| 8 | rs187613279 | 130915457 | G | A | - | - | 0.8148 | - | - |
| 9 | rs599452 | 22027402 | G | A | - | 0.3203 | - | - | - |
| 9 | rs1556515 | 22036367 | T | C | 0.2750 | - | 0.2201 | - | - |
| 9 | rs1412829 | 22043926 | A | G | - | - | - | 0.3531 | - |
| 9 | rs13285664 | 76915878 | C | T | 0.1892 | - | - | - | - |
| 9 | rs11143912 | 76924592 | C | A | - | 0.2272 | - | - | - |
| 10 | rs1408817 | 114498145 | G | A | - | - | 0.1515 | - | - |
| 11 | rs11233253 | 82398420 | T | G | - | 0.2090 | - | - | - |
| 11 | rs725271 | 95733346 | C | T | - | - | - | - | 0.5833 |
| 11 | rs7107785 | 95747337 | C | T | - | - | 0.1622 | - | - |
| 11 | rs648044 | 114030799 | G | A | - | - | 0.1691 | - | - |
| 11 | rs7125115 | 118478330 | A | G | - | - | - | - | 0.5011 |
| 11 | rs67307131 | 118480223 | C | T | 0.1492 | - | 0.3520 | - | - |
| 11 | rs11216998 | 118604837 | C | T | - | - | 0.2111 | - | - |
| 12 | rs6582308 | 76264083 | C | A | - | - | 0.1598 | - | - |

| Chr | Position (GRCh37) | rsID | Other Allele | Effect Allele | GWAS Effect Size | | | | |
|---|---|---|---|---|---|---|---|---|---|
| | | | | | All glioma | GBM | Non-GBM | IDH WT | IDH MUT |
| 14 | rs10131032 | 33250081 | A | G | - | - | 0.2987 | - | - |
| 15 | rs77387260 | 76537476 | C | T | 0.1833 | - | 0.3242 | - | - |
| 16 | rs11866537 | 90538 | C | T | - | 0.1934 | - | - | - |
| 16 | rs3751669 | 1035548 | G | A | - | - | 0.1793 | - | - |
| 16 | rs7186889 | 50092291 | G | A | - | 0.1633 | - | - | - |
| 16 | rs8051902 | 50099777 | C | T | 0.1307 | - | - | - | - |
| 17 | rs11870720 | 7434173 | C | T | 0.1672 | - | - | - | - |
| 17 | rs78378222 | 7571752 | T | G | 0.9269 | 0.9930 | 0.9490 | - | - |
| 20 | rs3746342 | 62289588 | C | A | 0.1817 | - | - | - | - |
| 20 | rs116668146 | 62308829 | A | G | 0.2445 | 0.2575 | - | - | - |
| 20 | rs2297440 | 62312299 | T | C | 0.2989 | 0.3732 | 0.1766 | - | - |
| 20 | rs6062492 | 62320242 | G | A | - | 0.2621 | - | - | - |
| 20 | rs6062302 | 62320968 | T | C | - | - | - | 0.4842 | - |
| 20 | rs116914291 | 62512441 | A | G | 0.3103 | - | - | - | - |
| 22 | rs2235573 | 38477930 | A | G | - | 0.1464 | - | - | - |
| 22 | rs2235264 | 38609950 | A | G | 0.1080 | - | - | - | - |

**Supplementary Table 3: Details of predictive performance of PRS-CT and PRS-CS in case-control analyses by histological subtype (all glioma, GBM, non-GBM).** Odds ratio (OR) per standard deviation increase in each standardized PRS. The "PRS model" column lists both the PRS construction method (PRS-CT or PRS-CS) and the training data phenotype (all glioma, GBM and non-GBM).

| Phenotype | Target data | PRS model | OR (95% CI) | P value | Nagelkerke's $R^2$ | Liability $R^2$ | AUC (95% CI) |
|---|---|---|---|---|---|---|---|
| All glioma | UK Biobank | PRS-CT (All glioma) | 1.64 (1.54, 1.76) | $4.1\times10^{-49}$ | 0.0191 | 0.0219 | 0.673 (0.653, 0.692) |
| | | PRS-CS (All glioma) | 1.70 (1.59, 1.82) | $5.7\times10^{-54}$ | 0.0214 | 0.0243 | 0.679 (0.660, 0.698) |
| | TCGA/WTCCC | PRS-CT (All glioma) | 1.44 (1.31, 1.58) | $4.5\times10^{-14}$ | 0.0233 | 0.00454 | - |
| | | PRS-CS (All glioma) | 1.53 (1.40, 1.69) | $1.1\times10^{-18}$ | 0.0323 | 0.00561 | - |
| GBM | UK Biobank | PRS-CT (GBM) | 1.83 (1.69, 1.98) | $9.4\times10^{-50}$ | 0.0269 | 0.0256 | 0.717 (0.695, 0.739) |
| | | PRS-CS (GBM) | 1.93 (1.78, 2.10) | $2.0\times10^{-54}$ | 0.0304 | 0.0282 | 0.724 (0.703, 0.746) |
| | TCGA/WTCCC | PRS-CT (GBM) | 1.46 (1.29, 1.65) | $2.2\times10^{-9}$ | 0.0218 | 0.00514 | - |
| | | PRS-CS (GBM) | 1.66 (1.46, 1.88) | $5.6\times10^{-15}$ | 0.0383 | 0.00880 | - |
| non-GBM | UK Biobank | PRS-CT (Non-GBM) | 1.66 (1.46, 1.88) | $4.6\times10^{-15}$ | 0.0202 | 0.0314 | 0.667 (0.620, 0.713) |
| | | PRS-CS (Non-GBM) | 1.75 (1.53, 2.00) | $1.7\times10^{-16}$ | 0.0232 | 0.0350 | 0.676 (0.630, 0.722) |
| | TCGA/WTCCC | PRS-CT (Non-GBM) | 1.67 (1.51, 1.86) | $6.8\times10^{-22}$ | 0.0528 | 0.0135 | - |
| | | PRS-CS (Non-GBM) | 1.74 (1.55, 1.96) | $6.5\times10^{-21}$ | 0.0530 | 0.0131 | - |

**Supplementary Table 4: Details of predictive performance of PRS-CT and PRS-CS for PRS trained with non-GBM data across non-GBM subtypes.** Odds ratio (OR) per standard deviation increase in each standardized PRS. Results not reported for oligoastrocytoma tumors in UK Biobank because model did not converge due to insufficient cases.

| Target phenotype | Target data | PRS model | OR (95% CI) | P value | Nagelkerke's $R^2$ | Liability $R^2$ | AUC (95% CI) |
|---|---|---|---|---|---|---|---|
| All non-GBM | UK Biobank (146 cases) | PRS-CT (Non-GBM) | 1.66 (1.46, 1.88) | $4.6\times10^{-15}$ | 0.0202 | 0.0314 | 0.667 (0.620, 0.713) |
| | | PRS-CS (Non-GBM) | 1.75 (1.53, 2.00) | $1.7\times10^{-16}$ | 0.0232 | 0.0350 | 0.676 (0.630, 0.722) |
| | TCGA/WTCCC (443 cases) | PRS-CT (Non-GBM) | 1.67 (1.51, 1.86) | $6.8\times10^{-22}$ | 0.0528 | 0.0135 | - |
| | | PRS-CS (Non-GBM) | 1.74 (1.55, 1.96) | $6.5\times10^{-21}$ | 0.0530 | 0.0131 | - |
| Astrocytoma | UK Biobank (95 cases) | PRS-CT (Non-GBM) | 1.41 (1.19, 1.68) | $6.8\times10^{-5}$ | 0.00805 | 0.0124 | 0.637 (0.580, 0.695) |
| | | PRS-CS (Non-GBM) | 1.43 (1.19, 1.70) | $9.2\times10^{-5}$ | 0.00799 | 0.0120 | 0.644 (0.589, 0.700) |
| | TCGA/WTCCC (168 cases) | PRS-CT (Non-GBM) | 1.45 (1.23, 1.70) | $7.8\times10^{-6}$ | 0.01990 | 0.00747 | - |
| | | PRS-CS (Non-GBM) | 1.51 (1.27, 1.80) | $2.9\times10^{-6}$ | 0.0231 | 0.00999 | - |
| Oligodendroglioma | UK Biobank (42 cases) | PRS-CT (Non-GBM) | 1.92 (1.55, 2.40) | $5.5\times10^{-9}$ | 0.0330 | 0.0611 | 0.758 (0.679, 0.836) |
| | | PRS-CS (Non-GBM) | 2.18 (1.73, 2.75) | $5.2\times10^{-11}$ | 0.0437 | 0.0788 | 0.755 (0.669, 0.842) |
| | TCGA/WTCCC (157 cases) | PRS-CT (Non-GBM) | 2.01 (1.73, 2.34) | $1.2\times10^{-19}$ | 0.0875 | 0.0350 | - |
| | | PRS-CS (Non-GBM) | 2.07 (1.74, 2.46) | $3.6\times10^{-16}$ | 0.0766 | 0.0285 | - |
| Oligoastrocytoma | UK Biobank (9 cases) | PRS-CT (Non-GBM) | - | - | - | - | - |
| | | PRS-CS (Non-GBM) | - | - | - | - | - |
| | TCGA/WTCCC (118 cases) | PRS-CT (Non-GBM) | 1.70 (1.42, 2.04) | $1.2\times10^{-8}$ | 0.0425 | 0.0207 | - |
| | | PRS-CS (Non-GBM) | 1.84 (1.50, 2.26) | $8.3\times10^{-9}$ | 0.0469 | 0.0218 | - |

**Supplementary Table 5: Details of predictive performance of PRS-CT and PRS-CS in case-control analyses by molecular subtype (*IDH* wildtype, *IDH* mutant).** Odds ratio (OR) per standard deviation increase in each standardized PRS. The PRS model column lists both the PRS construction method (PRS-CT or PRS-CS) and the training data phenotype (GBM, non-GBM, *IDH* wildtype, *IDH* mutant).

| Phenotype | Target data | PRS model | OR (95% CI) | P value | Nagelkerke's $R^2$ | Liability $R^2$ |
|---|---|---|---|---|---|---|
| *IDH* wildtype | TCGA/WTCCC | PRS-CT (*IDH* wildtype) | 1.51 (1.33, 1.72) | $4.0{\times}10^{-10}$ | 0.0246 | 0.00548 |
| | | PRS-CS (*IDH* wildtype) | 1.58 (1.40, 1.80) | $1.0{\times}10^{-12}$ | 0.0315 | 0.00701 |
| *IDH* wildtype | TCGA/WTCCC | PRS-CT (GBM) | 1.51 (1.34, 1.71) | $4.2{\times}10^{-11}$ | 0.0266 | 0.00629 |
| | | PRS-CS (GBM) | 1.72 (1.52, 1.95) | $2.3{\times}10^{-17}$ | 0.0453 | 0.0107 |
| *IDH* mutant | TCGA/WTCCC | PRS-CT (*IDH* mutant) | 1.81 (1.62, 2.03) | $2.5{\times}10^{-24}$ | 0.0658 | 0.0173 |
| | | PRS-CS (*IDH* mutant) | 1.85 (1.65, 2.07) | $2.5{\times}10^{-26}$ | 0.0772 | 0.0216 |
| *IDH* mutant | TCGA/WTCCC | PRS-CT (Non-GBM) | 1.85 (1.66, 2.06) | $9.5{\times}10^{-27}$ | 0.0751 | 0.0200 |
| | | PRS-CS (Non-GBM) | 1.94 (1.71, 2.20) | $2.4{\times}10^{-25}$ | 0.0729 | 0.0186 |

**Supplementary Table 6: Details of the odds ratio (OR) of each PRS category for PRS-CT and PRS-CS across histological and molecular subtypes.** The 40-60% category is used as the reference category, representing the "average" risk score in the general population. All PRS models are trained with subtype-specific GWAS discovery data corresponding to their target phenotype. For example, the *IDH* wildtype PRS is trained on summary statistics from a *IDH* wildtype GWAS.

| **Phenotype** | **Target data** | **PRS model** | **PRS category** | | | | |
|---|---|---|---|---|---|---|---|
| | | | 0-40% | 40-60% | 60-80% | 80-100% | 95-100% |
| All glioma | UK Biobank | PRS-CT | 0.64 (0.50-0.80) | 1 (Reference) | 1.40 (1.12-1.76) | 2.11 (1.71-2.61) | 3.31 (2.56-4.28) |
| | | PRS-CS | 0.61 (0.48-0.77) | 1 (Reference) | 1.29 (1.03-1.61) | 2.12 (1.72-2.60) | 3.22 (2.49-4.15) |
| | TCGA/WTCCC | PRS-CT | 0.68 (0.51- 0.90) | 1 (Reference) | 1.22 (0.91- 1.63) | 1.54 (1.16- 2.05) | 2.43 (1.66- 3.55) |
| | | PRS-CS | 0.67 (0.50- 0.90) | 1 (Reference) | 1.18 (0.87- 1.60) | 1.84 (1.39- 2.43) | 2.41 (1.64- 3.53) |
| GBM | UK Biobank | PRS-CT | 0.69 (0.52-0.93) | 1 (Reference) | 1.42 (1.06-1.90) | 2.82 (2.17-3.65) | 4.33 (3.18-5.90) |
| | | PRS-CS | 0.58 (0.43-0.77) | 1 (Reference) | 1.44 (1.09-1.90) | 2.55 (1.98-3.29) | 4.54 (3.39-6.10) |
| | TCGA/WTCCC | PRS-CT | 0.95 (0.63- 1.42) | 1 (Reference) | 1.65 (1.10- 2.49) | 2.16 (1.45- 3.21) | 3.27 (1.96- 5.45) |
| | | PRS-CS | 1.17 (0.76- 1.79) | 1 (Reference) | 1.85 (1.18- 2.90) | 3.61 (2.38- 5.49) | 4.24 (2.51- 7.17) |
| non-GBM | UK Biobank | PRS-CT | 0.75 (0.45-1.26) | 1 (Reference) | 1.13 (0.65-1.95) | 2.45 (1.53-3.94) | 4.64 (2.69-8.01) |
| | | PRS-CS | 0.64 (0.38-1.07) | 1 (Reference) | 1.00 (0.57-1.74) | 2.54 (1.60-4.04) | 4.76 (2.80-8.10) |
| | TCGA/WTCCC | PRS-CT | 0.71 (0.47- 1.06) | 1 (Reference) | 1.54 (1.02- 2.31) | 2.34 (1.60- 3.42) | 5.49 (3.48- 8.68) |
| | | PRS-CS | 0.68 (0.45- 1.04) | 1 (Reference) | 1.62 (1.07- 2.45) | 2.59 (1.77- 3.81) | 4.15 (2.58- 6.70) |
| *IDH* wildtype | TCGA/WTCCC | PRS-CT | 0.49 (0.33- 0.73) | 1 (Reference) | 0.75 (0.50- 1.13) | 1.71 (1.19- 2.46) | 2.26 (1.39- 3.66) |
| | | PRS-CS | 0.72 (0.48- 1.06) | 1 (Reference) | 1.20 (0.80- 1.81) | 2.31 (1.59- 3.35) | 2.85 (1.74- 4.66) |
| *IDH* mutant | TCGA/WTCCC | PRS-CT | 0.72 (0.45- 1.13) | 1 (Reference) | 1.00 (0.58- 1.74) | 2.94 (1.97- 4.39) | 5.64 (3.40- 9.34) |
| | | PRS-CS | 1.13 (0.72- 1.79) | 1 (Reference) | 1.41 (0.85- 2.34) | 4.01 (2.58- 6.22) | 8.05 (4.86-13.31) |

**Supplementary Table 7: PRS performance for logistic regression models that classify TCGA cases according to *IDH* mutation status.** Each row represents a different combination of covariates, which can include sex, age, first ten genetic ancestry principal components (PCs) and one of the four subtype-specific PRS models (GBM PRS, non-GBM PRS, *IDH* wildtype PRS, *IDH* mutant PRS). AUC and 95% confidence intervals (CIs) from 2,000 bootstrap samples are presented. The odds ratios (ORs) per standard deviation (SD) unit increase in PRS are provided for subtype discrimination models with only one risk score.

| PRS model | Covariates | | | | | | | OR | 95% CI | AUC | 95% CI |
|---|---|---|---|---|---|---|---|---|---|---|---|
| | **Sex** | **Age** | **Genetic PCs** | **GBM** | **non-GBM** | ***IDH* WT** | ***IDH* MUT** | | | | |
| None | X | X | | | | | | - | - | 0.839 | 0.808-0.867 |
| PRS-CT | | | X | X | | | | 0.76 | 0.66-0.88 | 0.632 | 0.592-0.669 |
| | | | X | | X | | | 1.83 | 1.57-2.13 | 0.713 | 0.679-0.750 |
| | | | X | X | X | | | - | - | 0.732 | 0.696-0.768 |
| | | | X | | | X | | 0.79 | 0.68-0.92 | 0.630 | 0.591-0.668 |
| | | | X | | | | X | 1.92 | 1.65-2.24 | 0.716 | 0.682-0.754 |
| | | | X | | | X | X | - | - | 0.722 | 0.686-0.757 |
| | | | X | X | X | X | X | - | - | 0.741 | 0.708-0.774 |
| | X | X | X | X | | | | 0.79 | 0.65-0.95 | 0.853 | 0.825-0.881 |
| | X | X | X | | X | | | 1.79 | 1.50-2.15 | 0.872 | 0.844-0.897 |
| | X | X | X | X | X | | | - | - | 0.879 | 0.854-0.904 |
| | X | X | X | | | X | | 0.80 | 0.66-0.97 | 0.853 | 0.824-0.879 |
| | X | X | X | | | | X | 1.95 | 1.62-2.37 | 0.875 | 0.849-0.900 |
| | X | X | X | | | X | X | - | - | 0.877 | 0.851-0.903 |
| | X | X | X | X | X | X | X | - | - | 0.882 | 0.857-0.907 |
| PRS-CS | | | X | X | | | | 0.60 | 0.52-0.70 | 0.674 | 0.636-0.711 |
| | | | X | | X | | | 1.69 | 1.46-1.96 | 0.691 | 0.653-0.726 |
| | | | X | X | X | | | - | - | 0.758 | 0.724-0.792 |
| | | | X | | | X | | 0.71 | 0.61-0.82 | 0.647 | 0.608-0.685 |
| | | | X | | | | X | 1.84 | 1.60-2.13 | 0.714 | 0.678-0.752 |
| | | | X | | | X | X | - | - | 0.734 | 0.699-0.770 |
| | | | X | X | X | X | X | - | - | 0.771 | 0.737-0.803 |
| | X | X | X | X | | | | 0.60 | 0.50-0.73 | 0.864 | 0.835-0.890 |
| | X | X | X | | X | | | 1.67 | 1.39-2.01 | 0.866 | 0.839-0.893 |
| | X | X | X | X | X | | | - | - | 0.890 | 0.865-0.913 |
| | X | X | X | | | X | | 0.70 | 0.57-0.85 | 0.857 | 0.829-0.884 |
| | X | X | X | | | | X | 1.87 | 1.56-2.26 | 0.874 | 0.848-0.899 |
| | X | X | X | | | X | X | - | - | 0.881 | 0.854-0.904 |
| | X | X | X | X | X | X | X | - | - | 0.895 | 0.872-0.918 |

**Supplementary Table 8: Estimated 5-year cumulative incidence at 60 years old stratified by sex, percentiles of the PRS distribution (PRS-CS) and histological subtype.** Low PRS corresponds to <20th percentile, average PRS is defined as >20th to <80th percentile and high PRS includes individuals in the >80th percentile of the normalized genetic risk score distributions. CI=confidence interval.

| Phenotype | Sex | Low PRS | | Average PRS | | High PRS | |
|---|---|---|---|---|---|---|---|
| | | Estimate | 95% CI | Estimate | 95% CI | Estimate | 95% CI |
| All glioma | Both | 0.039% | 0.030-0.051% | 0.083% | 0.074-0.094% | 0.174% | 0.151-0.200% |
| | Male | 0.052% | 0.039-0.068% | 0.110% | 0.097-0.126% | 0.231% | 0.198-0.268% |
| | Female | 0.031% | 0.023-0.040% | 0.066% | 0.057-0.076% | 0.137% | 0.116-0.161% |
| GBM | Both | 0.015% | 0.010-0.021% | 0.048% | 0.041-0.056% | 0.113% | 0.095-0.134% |
| | Male | 0.021% | 0.014-0.031% | 0.069% | 0.058-0.081% | 0.162% | 0.135-0.193% |
| | Female | 0.011% | 0.007-0.016% | 0.035% | 0.029-0.042% | 0.083% | 0.067-0.101% |
| Non-GBM | Both | 0.013% | 0.008-0.021% | 0.016% | 0.012-0.021% | 0.047% | 0.035-0.061% |
| | Male | 0.015% | 0.009-0.024% | 0.018% | 0.013-0.025% | 0.052% | 0.037-0.071% |
| | Female | 0.012% | 0.007-0.020% | 0.015% | 0.011-0.020% | 0.043% | 0.030-0.059% |

**Supplementary Table 9: Details of lifetime absolute risk by PRS category for PRS-CS across histological and molecular subtypes.** All PRS models are trained with subtype-specific GWAS discovery data corresponding to their target phenotype. For example, the *IDH* wildtype PRS is trained on summary statistics from a *IDH* wildtype GWAS.

| Phenotype | Target data | PRS category | | | | | |
|---|---|---|---|---|---|---|---|
| | | **0-20%** | **20-40%** | **40-60%** | **60-80%** | **80-100%** | **95-100%** |
| All glioma | UK Biobank | 0.23% (0.20%, 0.25%) | 0.35% (0.32%, 0.37%) | 0.46% (0.45%, 0.48%) | 0.61% (0.61%, 0.61%) | 1.00% (0.93%, 1.06%) | 1.39% (1.25%, 1.55%) |
| | TCGA/WTCCC | 0.27% (0.23%, 0.32%) | 0.39% (0.35%, 0.42%) | 0.48% (0.46%, 0.50%) | 0.61% (0.60%, 0.61%) | 0.90% (0.80%, 0.99%) | 1.18% (1.01%, 1.37%) |
| GBM | UK Biobank | 0.11% (0.09%, 0.12%) | 0.18% (0.16%, 0.20%) | 0.26% (0.24%, 0.27%) | 0.36% (0.36%, 0.37%) | 0.68% (0.63%, 0.73%) | 1.02% (0.90%, 1.16%) |
| | TCGA/WTCCC | 0.14% (0.11%, 0.18%) | 0.21% (0.19%, 0.24%) | 0.28% (0.26%, 0.30%) | 0.37% (0.36%, 0.37%) | 0.58% (0.51%, 0.66%) | 0.80% (0.64%, 0.99%) |
| Non-GBM | UK Biobank | 0.09% (0.07%, 0.11%) | 0.14% (0.12%, 0.15%) | 0.18% (0.17%, 0.19%) | 0.25% (0.24%, 0.25%) | 0.41% (0.36%, 0.47%) | 0.59% (0.48%, 0.72%) |
| | TCGA/WTCCC | 0.09% (0.07%, 0.11%) | 0.14% (0.12%, 0.15%) | 0.18% (0.17%, 0.19%) | 0.25% (0.24%, 0.25%) | 0.41% (0.36%, 0.46%) | 0.58% (0.48%, 0.70%) |
| *IDH* wildtype | TCGA/WTCCC | 0.16% (0.12%, 0.19%) | 0.23% (0.20%, 0.25%) | 0.29% (0.27%, 0.30%) | 0.37% (0.36%, 0.37%) | 0.56% (0.49%, 0.64%) | 0.76% (0.61%, 0.92%) |
| *IDH* mutant | TCGA/WTCCC | 0.08% (0.06%, 0.09%) | 0.12% (0.11%, 0.14%) | 0.17% (0.16%, 0.18%) | 0.24% (0.24%, 0.24%) | 0.43% (0.38%, 0.47%) | 0.63% (0.52%, 0.73%) |
